# Ultrasonic Texture Analysis Identifies Cardiac Microstructural Alterations in Immune-Inflammatory Disease

**DOI:** 10.1101/2020.06.10.20125443

**Authors:** Alan C. Kwan, Trevor Nguyen, Elizabeth H. Kim, Emmanuella Demosthenes, Gerran Salto, Eric Luong, Pranoti Hiremath, Debiao Li, Daniel S. Berman, Jignesh Patel, Susan Cheng

## Abstract

**Background:** Echocardiographic texture analysis using the signal intensity coefficient (SIC) can identify fibrosis-associated microstructural changes. This approach has not been tested in immune-inflammatory disease states such as myocarditis or transplant rejection.

**Methods:** Retrospective pilot analysis using the SIC was performed in populations including myocarditis (n=5), acute left anterior descending coronary artery STEMI (n=6), severe aortic stenosis with normal ejection fraction (n=7), ATTR amyloidosis (n=6), and cardiac transplant patients undergoing biopsy including patients with active rejection on histopathology (n=22), patients with history of rejection but no current rejection (n=15), and patients without history of or current rejection (n=5), and a healthy control group (n=28).

**Results:** Decreased SIC was noted in the myocarditis and transplant rejection populations, with decreasing SIC by presence and history of rejection. Consistent with prior literature, the SIC was elevated in cardiac conditions associated with fibrosis.

**Conclusions:** The SIC may be able to capture microstructural changes associated with immune-inflammatory processes such as cardiac transplant rejection and myocarditis.

## INTRODUCTION

Cardiac ultrasound, or echocardiography, is an essential first-line diagnostic tool that provides high-resolution macrostructural anatomical and functional information. However, it currently lacks sensitivity to identify myocardial microstructural changes which may precede macrostructural abnormalities.^1-3^ Identification of these changes would allow for earlier identification of at-risk patients and improved longitudinal phenotyping of disease states. We have previously described an echocardiographic texture analysis method called the signal intensity coefficient (SIC).^4^ The SIC provides quantitative information on myocardial microstructure by histogram-based radiomic analysis of intensities at the myocardial-pericardial interface.^5^ Elevated SIC has been associated with early hypertensive heart disease,^4^ preclinical myocardial alterations in metabolic syndrome,^6^ preclinical disease in genetic carriers of hypertrophic cardiomyopathy-associated genes,^7^ and incident heart failure.^8^ This method appears valid in fibrotic disease states, but has yet to be applied in immune-inflammatory disease states such as cardiac transplant rejection and myocarditis. In this pilot study we test the SIC across a broad range of populations including patients with immune-inflammatory myocardial conditions to attempt to identify echocardiographic signatures of microstructural changes across a broad spectrum of myocardial disease.

## METHODS

### Study Sample

Nine clinical populations were derived by medical record review. These groups included myocarditis (n=7), acute left anterior descending coronary artery (LAD) STEMI (n=6), severe aortic stenosis with normal ejection fraction (n=7), ATTR amyloidosis (n=6), and cardiac transplant patients undergoing biopsy including patients with active rejection on histopathology (n=22), patients with history of rejection but no current rejection (n=15), and patients without history of or current rejection (n=5), and a healthy control group (n=28). Myocarditis patients were defined by clinical documentation of acute myocarditis including fulminant viral myocarditis, giant cell myocarditis, or checkpoint inhibitor myocarditis. LAD STEMI required invasive coronary angiography, with echocardiography performed during the index hospitalization. ATTR amyloidosis was defined by patients with positive sodium pyrophosphate imaging and documentation of clinical suspicion. Severe aortic stenosis was defined by echocardiography and documentation of clinical suspicion. All echocardiography in cardiac transplant patients were performed on days of standard post-transplant biopsies for monitoring and diagnosis of cardiac transplant rejection. Biopsy status was determined by histological result of the biopsy performed on the same day (active rejection or not), and review of prior screening biopsy results back to the time of transplant. The control group consisted of patients referred for preoperative evaluation echocardiography with normal ejection fraction, no valvular abnormalities, and no history of chemotherapy. The institutional review board of Cedars-Sinai Medical Center approved all protocols related to the study.

### Echocardiographic Analysis

Quantitative echocardiographic analysis by signal intensity coefficient was performed as previously described.^8^ B-mode parasternal long axis views were manually assessed for visibility and appropriateness of analysis. DICOM image was imported into ImageJ (v 1.53, National Institutes of Health, Bethesda, MD). Region of interest (ROI) was applied to the myocardial-pericardial interface in the inferior wall during end diastole starting at the mitral leaflet tips. ImageJ macro was applied to a 5 x 30 ROI to identify intensity distribution by histogram. The high-spectrum signal intensity coefficient (HS-SIC) was calculated as the sum of 1-(p/256) where p is the 50^th^, 60^th^, 70^th^, 80^th^, and 90^th^ percentiles of signal intensity within the pericardial region of interest.

### Outcome of Interest

The primary outcome of interest was median HS-SIC within each demographic group with comparison between groups.

### Statistical Analysis

Kruskal-Wallis and Wilcoxon Rank Sum testing was used to compare groups. Trend testing in transplant patients was performed by testing fit to a linear model. R v4.0.0 and RStudio 1.2.5042 were used for statistical analysis. Significance was defined as p=0.05.

## RESULTS

Overall population and sub-population details are shown in **Table 1**. The overall mean age was 55.2±18.4 years old. Risk factors including diabetes, hypertension, smoking history, and known cardiac disease was frequently present, with hypertension in over half of the patients. Ejection fraction on average was normal, with significantly reduced ejection fraction seen in patients with myocarditis and STEMI. Number of patients in each group ranged between 5 in patient without active rejection or prior history of rejection to 28 in the control group.

**Table 1.** Population and demographics of subgroups. SIC: Signal Intensity Coefficient, STEMI: ST-Segment Elevation Myocardial Infarction, TAVR: Transcatheter Aortic Valve Replacement.

| Group | N | Mean Age | Male | T2DM | HTN | HL | Smoker | CAD | LVEF | High Spectrum SIC |
| --- | --- | --- | --- | --- | --- | --- | --- | --- | --- | --- |
| **Non-Cardiac Transplant Patients** |  |  |  |  |  |  |  |  |  |  |
| Controls | 28 | 51.1±16.9 | 12 (43%) | 3 (11%) | 11 (39%) | 7 (25%) | 4 (14%) | 2 (7%) | 65.5±3.71 | 0.545 (0.297-0.813) |
| Myocarditis | 7 | 53.5±20.7 | 2 (29%) | 1 (14%) | 2 (29%) | 1 (14%) | 1 (14%) | 0 (0%) | 33.7±15.3 | 0.340 (0.283-0.633) |
| STEMI | 6 | 52.8±20.6 | 5 (83%) | 0 (0%) | 3 (50%) | 3 (50%) | 0 (0%) | 6 (100%) | 42.7±17.1 | 0.582 (0.538-0.972) |
| ATTR Amyloidosis | 6 | 85.4±3.36 | 4 (67%) | 2 (33%) | 5 (83%) | 3 (50%) | 2 (33%) | 1 (17%) | 52.3±23.7 | 1.080 (0.616-1.13) |
| Pre-TAVR | 10 | 79.0±11.2 | 4 (40%) | 2 (20%) | 5 (50%) | 2 (20%) | 1 (10%) | 4 (40%) | 54.1±17.8 | 1.470 (0.729-1.78) |
| **Cardiac Transplant Recipients** |  |  |  |  |  |  |  |  |  |  |
| No Active, Negative History of Rejection | 5 | 49.6±14.1 | 5 (100%) | 3 (60%) | 3 (60%) | 3 (60%) | 1 (20%) | 1 (20%) | 55.8±1.92 | 0.656 (0.578-1.61) |
| No Active, Positive History of Rejection | 15 | 54.9±20.6 | 10 (67%) | 8 (53%) | 9 (60%) | 3 (20%) | 4 (27%) | 2 (13%) | 61.8±3.76 | 0.539 (0.398-0.705) |
| Active Rejection | 22 | 48.5±14.4 | 16 (73%) | 10 (45%) | 9 (41%) | 9 (41%) | 4 (18%) | 5 (23%) | 60.5±6.75 | 0.354 (0.209-0.614) |
| **Total** | 71 | 55.2±18.4 | 57 (80%) | 29 (41%) | 47 (66%) | 31 (44%) | 17 (24%) | 21 (30%) | 59.7±13.3 | 0.542 (0.335-0.864) |

\*T2DM, Type 2 diabetes; HM, hypertension; HL, hyperlipidemia; CAD, coronary artery disease; LVEF, left ventricular ejection fraction.

In non-transplant clinical patients, the median HS-SIC ranged from 0.340 in myocarditis patients to 1.47 in pre-TAVR aortic stenosis patients. In ascending order, the median values were myocarditis, STEMI, ATTR Amyloidosis, and pre-TAVR. Kruskal Wallace test for independent samples resulted in p=0.007. Within the transplant patients, patients who had no current and no history of rejection had the highest HS-SIC at 0.656, followed by those without current rejection but positive history of rejection with 0.539, and finally those with active/acute rejection on biopsy at 0.354. Kruskal Wallace test of independence resulted in p=0.041. Wilcoxon Rank sum test comparing patients without current or prior rejection versus those with active rejection had p=0.034. Trend testing for linearity in the transplant group was significant with p=0.018. The control group had a HS-SIC of 0.545, which was closest to patients with STEMI in the clinical population and patient with no active but positive history of rejection in the transplant population (**Figure 1**).

**Figure 1.**
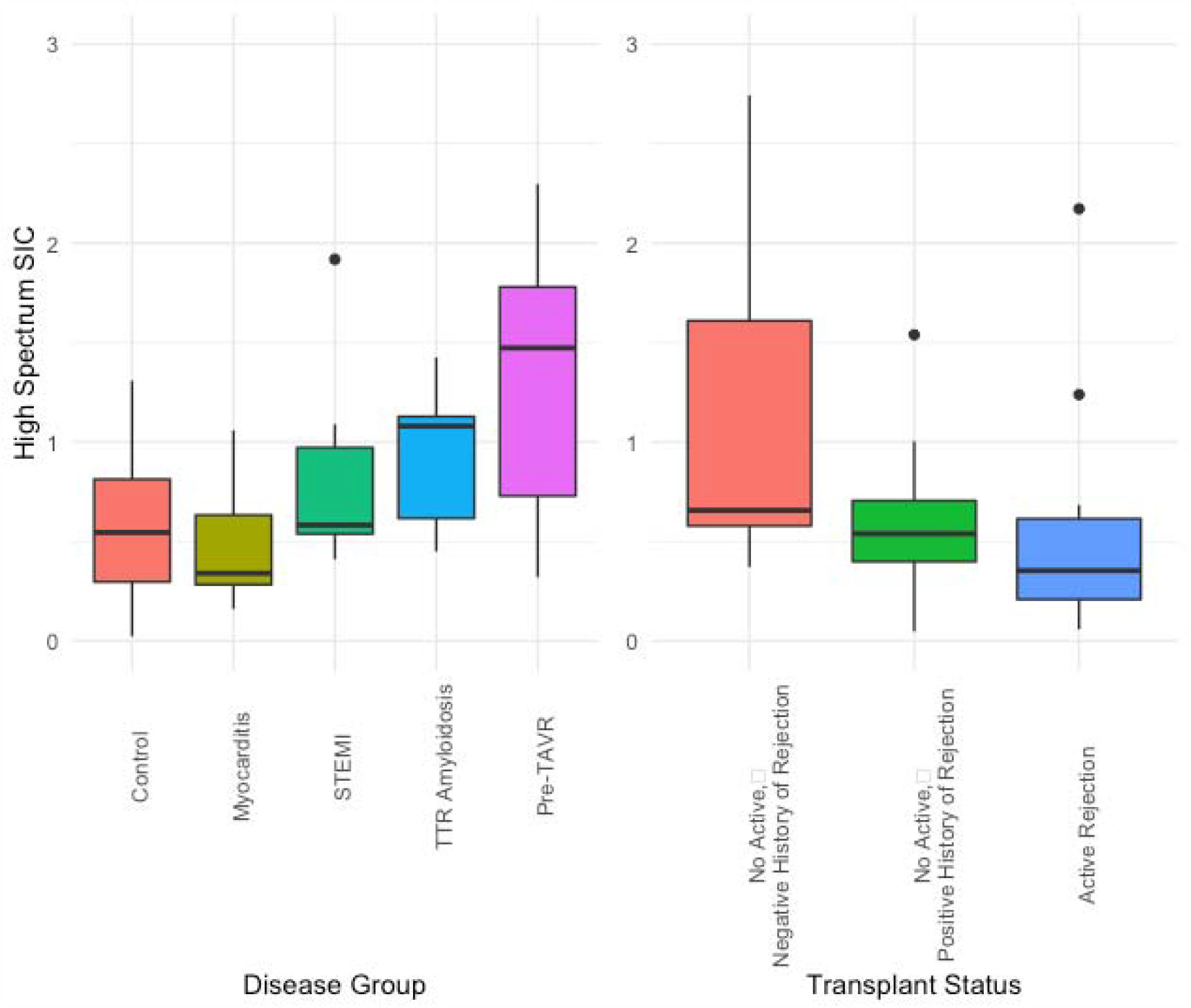
High Spectrum Signal Intensity Coefficient ranges for separate disease groups and post-cardiac transplant patients with no current / no history of rejection, no current / positive history of rejection, and current rejection. SIC: Signal Intensity Coefficient, STEMI: ST-Segment Elevation Myocardial Infarction, TAVR: Transcatheter Aortic Valve Replacement.

## DISCUSSION

In this pilot study, echocardiographic texture analysis was performed on small populations of patients with varying clinical cardiac conditions. This included standard clinical cardiac conditions as well as a population of patients who had received cardiac transplantation and had varying degrees of current or historical rejection. While these groups were low in number, certain characteristics emerged. As expected, conditions associated with myocardial fibrosis, such as aortic stenosis and amyloidosis had elevated HS-SIC values. Less expected was the observation that patients with immune-inflammatory conditions such as myocarditis or active transplant rejection had values had the lowest values. Within the transplant population, the three groups of no current and no history of rejection, no current but positive history of rejection, and current rejection showed a stepwise decrease in HS-SIC values consistent with the expected degree of immune-inflammatory activity in these groups.

Elevated SIC has been previously associated with microstructural changes in diseases associated with cardiac fibrosis including hypertension, obesity, hypertrophic cardiomyopathy, and incident congestive heart failure. This methodology has not previously been tested in immune-inflammatory conditions such as myocarditis and transplant rejection. Initial expectation with transplant rejection was cases may show higher degrees of fibrosis and therefore elevated HS-SIC values, the degree of rejection appears to be associated with a decrease in HS-SIC. The similar reduction in HS-SIC in the myocarditis population gives us further support that this may be a true signal despite our pilot population sizes.

There are multiple limitations with this pilot study, most notably the small and uneven population sizes. Associated clinical risk factors varied widely between populations due to the small sample size. A clearer signal with the ability to control for risk factors will be feasible with a larger population. Additionally, while the HS-SIC has been studied in fibrosis, we lack a priori histological associations in immune-inflammatory disease, and therefore our observations of decreased values are only associative at this time. Further investigation is required.

In conclusion, the HS-SIC demonstrates variation across different clinical populations in both standard and post-cardiac-transplant patients. These variations appear to correlate with expected histological characteristics. Specifically, populations with higher HS-SIC are populations where we would expect increased myocardial fibrosis. Our study presents new data that patients with lower HS-SIC may have increased degrees of immune-inflammatory disease. Further investigation of patients with immune-inflammatory disease is required.

## Disclosures

The authors have no relevant disclosures

## Data Availability

De−identified data are available upon request and in accordance with standard institutional data sharing agreements.

## Sources of Funding

This work was supported by the National Institutes of Health contract N01-HC-25195 HHSN268201500001I and grants T32HL116273, R01 HL 077477, R01HL131532, R01HL134168, R01 DK080739, R01HL126136, R01 HL 080124, R01 HL 077477, and R01 HL 70100; the Barbra Streisand Women’s Cardiovascular Research and Education Program; and, the Erika Glazer Women’s Heart Health Project.

## Notes

### Competing Interest Statement

The authors have declared no competing interest.

### Author Declarations

The institutional review board of Cedars-Sinai Medical Center approved all protocols.

